# Ipsilateral rest tremor-dopamine transporter correlation reflects a broader dopaminergic difference, not tremor-specific pathophysiology

**DOI:** 10.64898/2026.08.24.26361220

**Authors:** Marcelo D Mendonça, Joaquim Alves da Silva

## Abstract

**Background and Objectives:** To test whether the positive correlation between rest tremor (RT) and ipsilateral striatal DAT binding reflects a tremor-specific ipsilateral mechanism or simply the globally better-preserved dopamine terminals in RT patients.

**Methods:** We compared ipsilateral and contralateral correlations between striatal binding, rest tremor, bradykinesia and rigidity in two cross-sectional Parkinson disease cohorts from the Parkinson’s Progression Markers Initiative (baseline, N=1055; follow-up, median 2.2 years, N=652). We tested whether correlations survived permutation testing that shuffled severity scores isolating severity-dependent effects from group-level effects and used out-of-sample prediction to compare how well binding predicted symptom presence versus severity.

**Results:** Bradykinesia and rigidity showed large contralateral correlations, distinguishable from the permutation null in every comparison, and predicted both presence and severity. Rest tremor’s ipsilateral correlation was not distinguishable from the null in most comparisons. Striatal binding, both ipsi or contralateral, predicted its presence (AUC 0.55–0.61) but not its severity (R^2^ ≤0.002), in both cohorts.

**Conclusions:** Our findings argue against a direct pathophysiologic link between tremor amplitude and ipsilateral dopaminergic function. More parsimoniously, the ipsilateral binding–RT correlation likely reflects a distinct degeneration pattern in patients with RT rather than a graded, dose-dependent circuit mechanism.

## Introduction

The relationship between motor symptoms and striatal dopamine transporter (DAT) binding is central to how molecular imaging is used to understand Parkinson disease (PD). Yet a correlation between a clinical sign and a biomarker can arise from more than one process: it may reflect the sign’s own pathophysiology, or a broader feature of disease architecture associated with it without causing it. Distinguishing between these possibilities matters to develop a pathophysiological understanding.

Tremor and DAT binding illustrate this problem well. An unexpected positive correlation between tremor and ipsilateral striatal binding has been observed repeatedly, in several early, underpowered cohorts^1,2^, in our own prior work (most prominent in caudate)^3^, and with a distinct tracer^4^. Niemi and colleagues reported the same pattern in two independent samples^5,6^ concluding that it “primarily support[s] a pathophysiologic link between tremor amplitude and dopaminergic function.”^6^ The correlation itself is a robust, reproducible finding, replicated across cohorts, tracers, and groups. What remains open is its interpretation.

One hypothesis holds that it reflects a direct, tremor-specific, dose-dependent relationship between local dopaminergic integrity and ipsilateral tremor amplitude. An alternative, which we proposed using *in silico* modelling^3,7^ and which was subsequently discussed but not resolved^8^, holds that it can emerge without any true ipsilateral tremor amplitude-DAT relationship, as tremor emerges from processes that are associated with a globally less pronounced striatal DAT binding impairment, for reasons unrelated to tremor. Since simulation alone couldn’t adjudicate between these accounts, we tested them directly here.

The two hypotheses make different predictions: a direct mechanism should predict tremor severity, not only presence, whereas a non-specific group-level difference should predict presence but not severity. We tested this with a permutation test asking whether the correlation survives shuffling severity among tremor-positive patients, and with out-of-sample prediction comparing how well binding predicts severity versus symptom presence.

## Methods

We used two cross-sectional PD cohorts from the PPMI: a baseline cohort (screening/baseline visit, N=1055) and a cohort built from non-baseline visits (median 2.2 years post-enrollment, N=652). Each patient was matched to an Off-state MDS-UPDRS-III assessment performed within 91 days of DAT-SPECT. Imaging data were obtained from PPMI’s centrally computed DAT-SPECT analysis, using caudate and putamen specific binding ratios (CBR, PBR) derived from the pipeline’s standardized quantification. For rest tremor, bradykinesia, and rigidity, we derived composite left- and right-sided severity scores from individual MDS-UPDRS-III items. For each symptom, side, cohort, and region, we compared the observed Spearman correlation between severity and ipsilateral or contralateral binding against a null distribution generated by holding symptom-absent patients fixed and randomly permuting severity values among symptom-positive patients (10,000 permutations, two-sided). Statistical significance was defined at α=0.05, with Benjamini-Hochberg false discovery rate (FDR) procedure for multiple comparisons.

We additionally tested how well unilateral striatal binding predicted symptom status on the same (ipsilateral) and opposite (contralateral) side. Presence (symptom score >0) was modelled by leave-one-out cross-validated logistic regression and evaluated by area under the curve (AUC); significance was assessed by Mann-Whitney U test on the binding values. Where leave-one-out predictions were numerically unstable, AUC was substituted with a rank-based estimate derived directly from the binding values. Severity among symptom-positive patients was modelled by leave-one-out cross-validated linear regression and evaluated by R^2^; significance was assessed by Pearson correlation on the non-cross-validated values.

## Results

On the baseline cohort (63.5 ± 9.5 years, MDS-UPDRS III: 22.9 ± 11.0) bradykinesia and rigidity showed, as expected, large contralateral correlations with striatal binding that were far outside the permutation null in caudate and putamen (ρ –0.20 to –0.38; all p<0.05, Figure 1A-B and Table 1), with negligible ipsilateral correlations (Table 1). Rest tremor showed a different pattern instead: modest positive ipsilateral correlations that are within the null distribution (ρ +0.11 to +0.22, all p>0.05, Figure 1C and Table 1), with modest negative contralateral correlations. Results were similar on the follow-up cohort (Table 1, 65.6 ± 9.2 years, MDS-UPDRS III: 27.8 ± 12.5, Median time since enrollment: 2.2 years, IQR: 2.0-4.0 years).

**Table 1.**
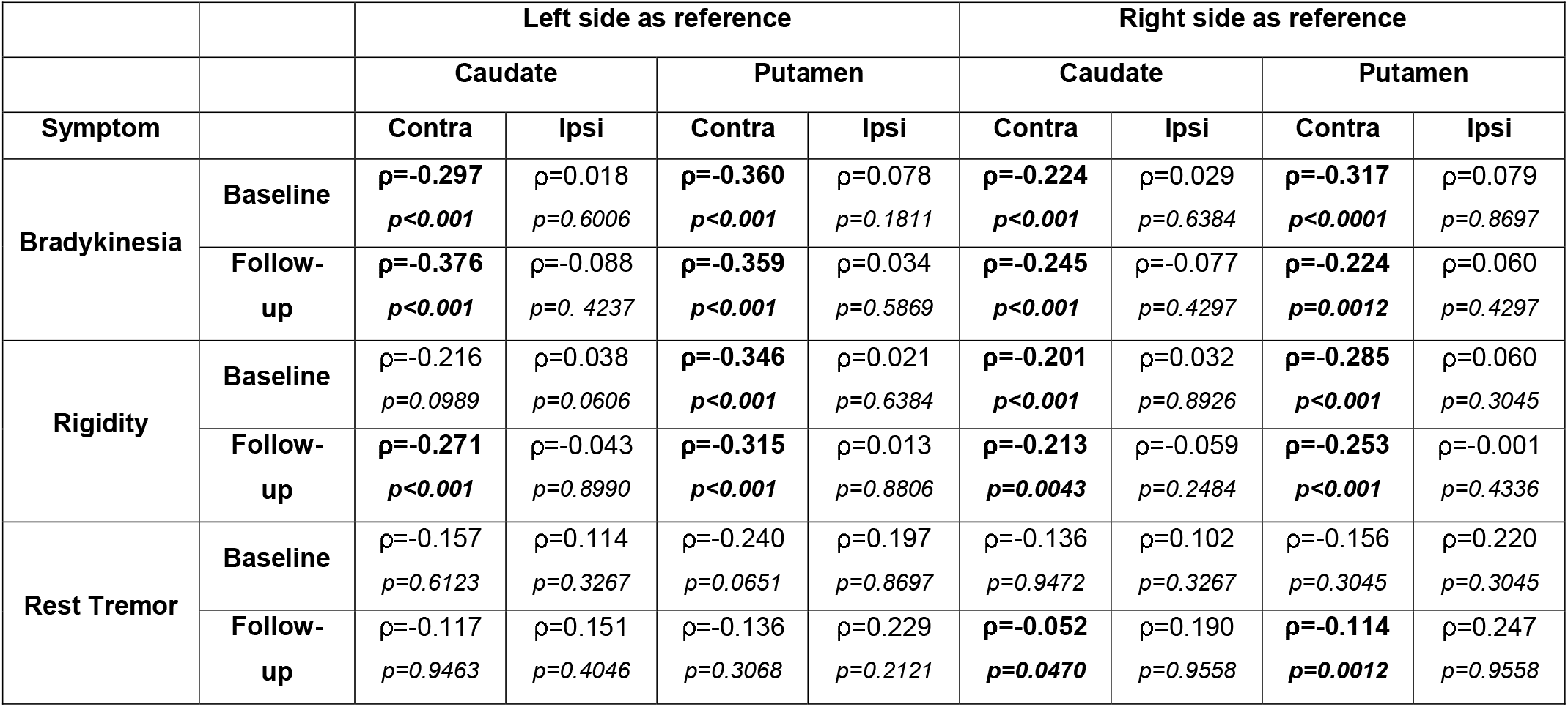
Results of the permutation test for bradykinesia, rigidity, and rest tremor, baseline and follow-up cohorts. For each symptom, left and right sides were each used as an independent reference side, against which the opposite (contralateral) and same (ipsilateral) side’s striatal binding were compared. After correction for multiple comparisons, significant effects are concentrated on the contralateral side for bradykinesia and rigidity, with consistently stronger effects in putamen than caudate. Rest tremor’s ipsilateral correlation was not distinguishable from the permutation null in any comparison, despite modest positive point estimates. A contralateral correlation for rest tremor did reach significance in the follow-up cohort only (both caudate and putamen, right-side reference). ρ = observed Spearman correlation between symptom severity and striatal binding; p = Benjamini-Hochberg false discovery rate-adjusted p-value

|  |  | Left side as reference |  |  |  | Right side as reference |  |  |  |
| --- | --- | --- | --- | --- | --- | --- | --- | --- | --- |
|  |  | Caudate |  | Putamen |  | Caudate |  | Putamen |  |
| Symptom |  | Contra | Ipsi | Contra | Ipsi | Contra | Ipsi | Contra | Ipsi |
| Bradykinesia | Baseline | $\rho=-0.297$<br>$p<0.001$ | $\rho=0.018$<br>$p=0.6006$ | $\rho=-0.360$<br>$p<0.001$ | $\rho=0.078$<br>$p=0.1811$ | $\rho=-0.224$<br>$p<0.001$ | $\rho=0.029$<br>$p=0.6384$ | $\rho=-0.317$<br>$p<0.0001$ | $\rho=0.079$<br>$p=0.8697$ |
| | Follow-up | $\rho=-0.376$<br>$p<0.001$ | $\rho=-0.088$<br>$p=0.4237$ | $\rho=-0.359$<br>$p<0.001$ | $\rho=0.034$<br>$p=0.5869$ | $\rho=-0.245$<br>$p<0.001$ | $\rho=-0.077$<br>$p=0.4297$ | $\rho=-0.224$<br>$p=0.0012$ | $\rho=0.060$<br>$p=0.4297$ |
| Rigidity | Baseline | $\rho=-0.216$<br>$p=0.0989$ | $\rho=0.038$<br>$p=0.0606$ | $\rho=-0.346$<br>$p<0.001$ | $\rho=0.021$<br>$p=0.6384$ | $\rho=-0.201$<br>$p<0.001$ | $\rho=0.032$<br>$p=0.8926$ | $\rho=-0.285$<br>$p<0.001$ | $\rho=0.060$<br>$p=0.3045$ |
| | Follow-up | $\rho=-0.271$<br>$p<0.001$ | $\rho=-0.043$<br>$p=0.8990$ | $\rho=-0.315$<br>$p<0.001$ | $\rho=0.013$<br>$p=0.8806$ | $\rho=-0.213$<br>$p=0.0043$ | $\rho=-0.059$<br>$p=0.2484$ | $\rho=-0.253$<br>$p<0.001$ | $\rho=-0.001$<br>$p=0.4336$ |
| Rest Tremor | Baseline | $\rho=-0.157$<br>$p=0.6123$ | $\rho=0.114$<br>$p=0.3267$ | $\rho=-0.240$<br>$p=0.0651$ | $\rho=0.197$<br>$p=0.8697$ | $\rho=-0.136$<br>$p=0.9472$ | $\rho=0.102$<br>$p=0.3267$ | $\rho=-0.156$<br>$p=0.3045$ | $\rho=0.220$<br>$p=0.3045$ |
| | Follow-up | $\rho=-0.117$<br>$p=0.9463$ | $\rho=0.151$<br>$p=0.4046$ | $\rho=-0.136$<br>$p=0.3068$ | $\rho=0.229$<br>$p=0.2121$ | $\rho=-0.052$<br>$p=0.0470$ | $\rho=0.190$<br>$p=0.9558$ | $\rho=-0.114$<br>$p=0.0012$ | $\rho=0.247$<br>$p=0.9558$ |

**Figure 1.**
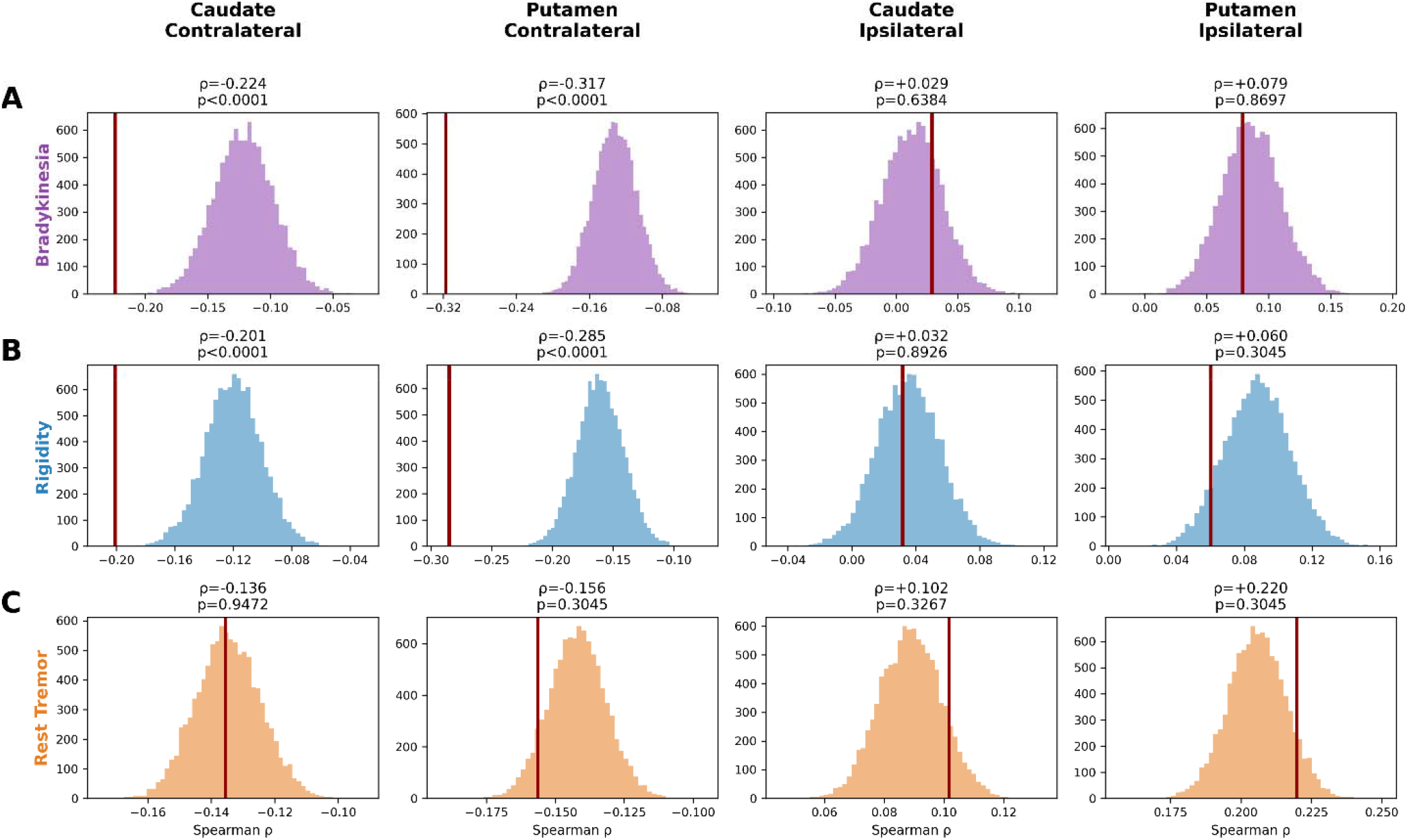
Permutation (shuffle-control) test for bradykinesia (A), rigidity (B), and rest tremor (C), baseline cohort (N=1055). For each symptom, the right side was used as the reference side: “ipsilateral” refers to right-side striatal binding relative to right-sided symptom severity, and “contralateral” refers to left-side striatal binding relative to the same right-sided symptom. The observed Spearman correlation (red line) is shown against a null distribution built by holding symptom-absent patients fixed and randomly permuting severity values among symptom-positive patients 10,000 times. p-values are Benjamini-Hochberg FDR-adjusted, computed across all 24 tests in this cohort. Bradykinesia (A) and rigidity (B) show large, highly significant contralateral correlations (stronger in putamen than in caudate) with negligible ipsilateral correlations; rest tremor (C) shows a distinct pattern: modest ipsilateral correlations, not distinguishable from the permutation null.

Unlike bradykinesia’s and rigidity’s contralateral correlations, rest tremor’s ipsilateral correlation was significantly different from zero by conventional testing, yet not distinguishable from the permutation null in most comparisons (Table 1). This is consistent with a group-level difference in striatal binding rather than a graded, severity-dependent relationship.

We tested this further using out-of-sample prediction of presence and severity.

Using unilateral caudate binding to predict contralateral and ipsilateral symptom status directly, rest tremor’s presence was predicted from binding on either side, in 7 of 8 comparisons (Figure 2A, logistic regression AUC 0.55–0.61; p<0.01), with no contralateral advantage. Its severity among affected patients, by contrast, was not predicted by any comparison (R^2^ ≤0.002). Tremor presence was predictable, but severity not, supporting a dopaminergic phenotype rather than a dose-dependent effect. By contrast bradykinesia showed the opposite pattern: contralateral putamen binding predicted presence (Figure 2B, AUC 0.65–0.74, p<0.001 in all comparisons) and severity (R^2^ 0.015–0.056, p<0.001 in all comparisons), whereas ipsilateral binding predicted neither severity (R^2^ ≤0) nor presence (1 of 8 comparisons). This pattern is the one expected for a direct, local, dose-dependent dopaminergic mechanism. Results for the alternate region (putamen for tremor, caudate for bradykinesia) were comparable (eFigure1).

**Figure 2.**
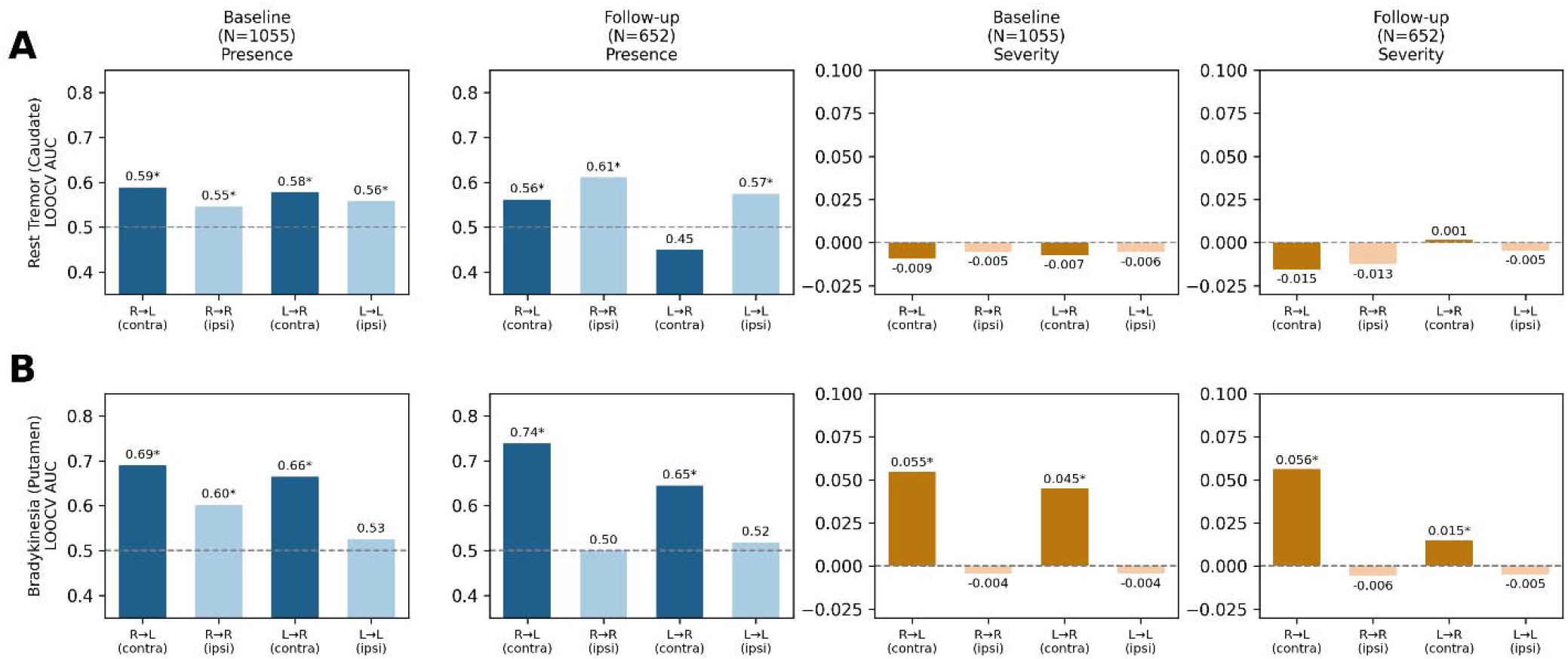
Out-of-sample prediction of symptom presence and severity from unilateral striatal binding, rest tremor (caudate, A) and bradykinesia (putamen, B), baseline and follow-up cohorts. For each symptom, both the right and left side were used as the reference side in turn, giving four reference-side/target-side combinations per panel: right-side binding predicting left-sided (contralateral) and right-sided (ipsilateral) symptom status, and left-side binding predicting right-sided (contralateral) and left-sided (ipsilateral) symptom status. Presence (symptom score >0) was modelled by leave-one-out cross-validated logistic regression (AUC); severity among symptom-positive patients was modelled by leave-one-out cross-validated linear regression (R^2^). Asterisks indicate significance after Benjamini-Hochberg correction for multiple comparisons (p<0.05), applied separately across the 16 presence tests and the 16 severity tests shown. Bradykinesia’s (B) contralateral binding predicts both presence and severity at both timepoints; its ipsilateral binding predicts neither. Rest tremor’s (A) presence is predicted from binding on either side, with no consistent contralateral advantage, but its severity is not predicted in any comparison.

## Discussion

Our findings support a dopaminergic phenotype associated with the presence of rest tremor, but not a dose-dependent relationship between striatal dopaminergic loss and tremor severity. Across both permutation testing and out-of-sample modelling, striatal binding predicted whether rest tremor was present, in both ipsilateral and contralateral channels alike, but carried no information about its severity. Bradykinesia and rigidity, by contrast, showed the expected contralateral relationship with striatal binding, confirming our methods could detect a genuine association where one exists. The absence of an equivalent relationship for tremor severity therefore argues against striatal dopaminergic dysfunction as a direct determinant of tremor amplitude.

This distinction reconciles our findings with prior literature. Rest tremor’s ipsilateral correlation was significantly different from zero in both cohorts, consistent with prior reports, tracers, and groups. Our permutation test was more demanding: shuffling severity among tremor-positive patients preserved presence while abolishing the severity gradient, and we found no evidence that binding tracks severity beyond presence. Prior reports were not wrong that a correlation exists: they could not distinguish a case-mix-driven correlation from a graded circuit mechanism, a distinction that matters for mechanistic interpretation.

This study has several limitations. Symptom severity was derived from ordinal MDS-UPDRS-III item scores, which coarsen severity and may limit sensitivity to a real but small dose-response relationship. Nevertheless, the same measures detected the expected relationship for bradykinesia and rigidity. Bradykinesia’s composite score is derived from five items with a wider dynamic range than rest tremor’s and is unilaterally present in most patients. Bradykinesia’s presence results in Figure 2 should therefore be interpreted cautiously; the severity analysis, unaffected by this threshold, is the more informative comparison. Finally, our sample is drawn from a single multi-site cohort (PPMI), and replication in independent cohorts would strengthen confidence.

Previous findings showed that patients with rest tremor differ in their pattern of striatal dopamine transporter binding^3,9,10^, and this difference alone is sufficient to produce a modest ipsilateral correlation^3,7^. Our findings provide further evidence for this non-lateralized dopaminergic phenotype in rest tremor. This is a more parsimonious explanation than one requiring an ipsilateral circuit, a logic at odds with the fundamentally contralateral organization of motor control. This is also consistent with long-standing observations that tremor-dominant PD follows a distinct clinical course^11,12^, and with the entity of benign tremulous parkinsonism^13,14^. Whether this reflects a genuinely different pattern of nigrostriatal degeneration^15^, or an upstream factor independently influencing both tremor liability and dopaminergic vulnerability, remains to be determined.

## Supporting information

eFigure1

## Data Availability

Data used in the preparation of this article were obtained on August, 2, 2026 from the Parkinson's Progression Markers Initiative (PPMI) database (https://www.ppmi-info.org/access-data-specimens/download-data), RRID:SCR_006431.

## Acknowledgement and Data Availability Statement

Data used in the preparation of this article were obtained on August, 2, 2026 from the Parkinson’s Progression Markers Initiative (PPMI) database (https://www.ppmi-info.org/access-data-specimens/download-data), RRID:SCR_006431. For up-to-date information on the study, visit http://www.ppmi-info.org. PPMI – a public-private partnership – is funded by the Michael J. Fox Foundation for Parkinson’s Research and funding partners, including 4D Pharma, Abbvie, AcureX, Allergan, Amathus Therapeutics, Aligning Science Across Parkinson’s, AskBio, Avid Radiopharmaceuticals, BIAL, BioArctic, Biogen, Biohaven, BioLegend, BlueRock Therapeutics, Bristol-Myers Squibb, Calico Labs, Capsida Biotherapeutics, Celgene, Cerevel Therapeutics, Coave Therapeutics, DaCapo Brainscience, Denali, Edmond J. Safra Foundation, Eli Lilly, Gain Therapeutics, GE HealthCare, Genentech, GSK, Golub Capital, Handl Therapeutics, Insitro, Jazz Pharmaceuticals, Johnson & Johnson Innovative Medicine, Lundbeck, Merck, Meso Scale Discovery, Mission Therapeutics, Neurocrine Biosciences, Neuron23, Neuropore, Pfizer, Piramal, Prevail Therapeutics, Roche, Sanofi, Servier, Sun Pharma Advanced Research Company, Takeda, Teva, UCB, Vanqua Bio, Verily, Voyager Therapeutics, the Weston Family Foundation and Yumanity Therapeutics.

The scripts used for analyses are publicly available in the following GitHub repository: mdmendonca/IpsilateralTremor.

This study is a secondary analysis of de-identified data from PPMI, a multi-site observational study approved by the institutional review board at each participating site; all PPMI participants provided written informed consent at enrollment. No additional consent was required for this secondary analysis.

Claude (Antrophic; Sonnet 5) was used to assist with refinement of the manuscript text and with coding during the analyses. All AI-assisted text and code were reviewed and verified by the authors, including line-by-line review of the publicly available analysis code. The authors take full responsibility for the final manuscript, analyses, and interpretation.

