## Supplementary material for "Ipsilateral rest tremor-dopamine transporter correlation reflects a broader dopaminergic difference, not tremor-specific pathophysiology": eFigure1

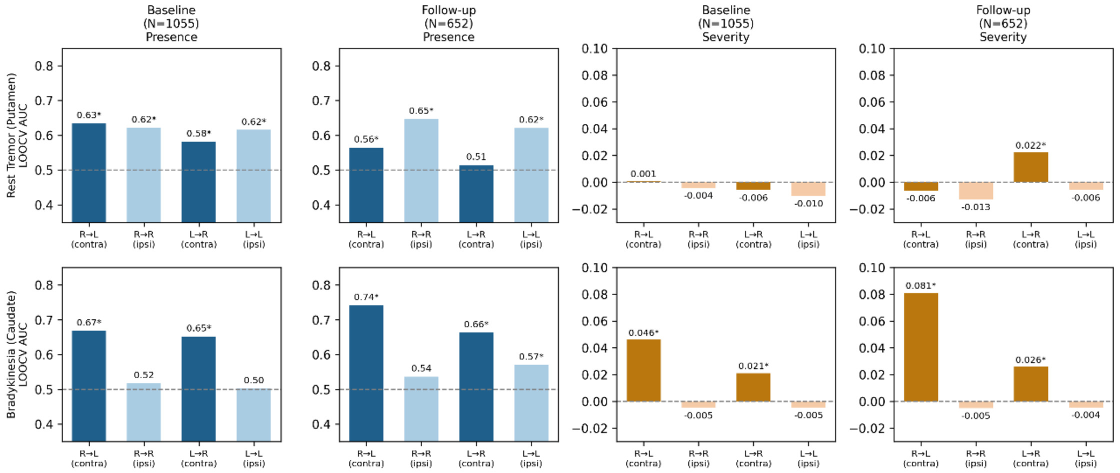


**Supplementary Figure 1. Out-of-sample prediction of symptom presence and severity from unilateral striatal binding, using the alternate region for each symptom: rest tremor (putamen) and bradykinesia (caudate), baseline and follow-up cohorts.** This figure repeats the analysis shown in Figure 2 using the region not shown there for each symptom, as a robustness check. For each symptom, both the right and left side were used as the reference side in turn, giving four reference-side/target-side combinations per panel: right-side binding predicting left-sided (contralateral) and right-sided (ipsilateral) symptom status, and left-side binding predicting right-sided (contralateral) and left-sided (ipsilateral) symptom status. Presence (symptom score >0) was modelled by leave-one-out cross-validated logistic regression (AUC); severity among symptom-positive patients was modelled by leave-one-out cross-validated linear regression ($R^2$). Asterisks indicate significance after Benjamini-Hochberg correction for multiple comparisons ($p<0.05$), applied separately across the 16 presence tests and the 16 severity tests shown. Results are directionally consistent with Figure 2: bradykinesia's contralateral caudate binding predicts both presence and severity at both timepoints, while its ipsilateral binding predicts neither. Rest tremor's presence is again predicted from putamen binding on either side.
